# Sensitivity and consistency of long- and short-read metagenomics and epicPCR for the detection of antibiotic resistance genes and their bacterial hosts in wastewater

**DOI:** 10.1101/2023.08.08.23293828

**Authors:** Esther G. Lou, Yilei Fu, Qi Wang, Todd J. Treangen, Lauren B. Stadler

**Affiliations:** Department of Civil and Environmental Engineering, Rice University, 6100 Main Street, Houston, TX 77005, USA; Department of Computer Science, Rice University, 6100 Main Street, Houston, TX 77005, USA

**Author notes:** These two authors contributed equally to this work (Esther G. Lou, Yilei Fu).

**Keywords:** antibiotic resistance genes (ARGs), antibiotic resistant bacteria (ARB), wastewater treatment plant (WWTP), metagenomics, epicPCR

## Abstract

Wastewater surveillance is a powerful tool to assess the risks associated with antibiotic resistance in communities. One challenge is selecting which analytical tool to deploy to measure risk indicators, such as antibiotic resistance genes (ARGs) and their respective bacterial hosts. Although metagenomics is frequently used for analyzing ARGs, few studies have compared the performance of long-read and short-read metagenomics in identifying which bacteria harbor ARGs in wastewater. Furthermore, for ARG host detection, untargeted metagenomics has not been compared to targeted methods such as epicPCR. Here, we 1) evaluated long-read and short-read metagenomics as well as epicPCR for detecting ARG hosts in wastewater, and 2) investigated the host range of ARGs across the WWTP to evaluate host proliferation. Results highlighted long-read revealed a wider range of ARG hosts compared to short-read metagenomics. Nonetheless, the ARG host range detected by long-read metagenomics only represented a subset of the hosts detected by epicPCR. The ARG-host linkages across the influent and effluent of the WWTP were characterized. Results showed the ARG-host phylum linkages were relatively consistent across the WWTP, whereas new ARG-host species linkages appeared in the WWTP effluent. The ARG-host linkages of several clinically relevant species found in the effluent were identified.

## 1. Introduction

The worldwide propagation and dissemination of antibiotic resistance have raised serious public health concerns. An estimated 1.27 million deaths were attributed to bacterial antibiotic resistant infections in 2019^1^. To mitigate this risk, a comprehensive understanding of antibiotic resistance in humans, animals, and the environment is needed^2^. Wastewater treatment plants (WWTPs) are regarded as hotspots of antibiotic resistance in the environment^3,4^, and their role in the dissemination of antibiotic resistance is complex. Wastewater treatment processes generally remove antibiotic resistance genes (ARGs) and antibiotic resistant bacteria (ARB) from sewage through different mechanisms such as, anaerobic^5,6^ and aerobic processes^7,8^, coagulation and sedimentation^9^, membrane filtration^10,11^, and disinfection^12,13^. In spite of the significant removal of the overall abundance of ARGs and ARB by WWTPs^14–16^, certain ARGs and ARB can be persistent or even enriched across the treatment units^17–19^, representing a major source of ARGs and ARBs in receiving waterbodies. For example, ARGs and microorganisms discharged by WWTPs can persist in receiving-river biofilms^20^ and sediments^21,22^. Furthermore, ARGs and ARB after establishing in the effluent-receiving environment can propagate to distant areas away from the discharge point^21,23^.

Previous studies have proposed metrics for evaluating risks associated with ARGs in the environment to public health^24,25^. One key component is identification of the bacterial host of the ARG, as a pathogenic bacteria harboring an ARG is a far greater public health risk as compared to a non-pathogenic environmental bacterial host of the same ARG^26,27^. A second component is whether an ARG is associated with mobile genetic elements (MGEs), which can directly and indirectly mediate horizontal gene transfer (HGT) of ARGs among microorganisms^28–30^. Thus, methods are needed that not only identify ARGs in environmental samples, but also provide information that link those ARGs to their microbial hosts and contextual information about whether the ARGs are associated with MGEs.

One of the most widely used methods to analyze antibiotic resistance in wastewater is metagenomic sequencing^31–33^. Next-generation sequencing (i.e., short-read) coupled with *de novo* assembly recovers ARG-host and ARG-MGE linkages by screening taxonomical markers and MGEs on the assembled ARG-carrying contigs^29,34,35^. However, this method suffers from limited detection sensitivity due to the low percentage of raw reads mapping back to the assembled contigs^36^ and intergenomic assembly errors^37^. Importantly, a large portion of reads associated with MGEs fail to assemble because of the extended homologous and mosaic sequences found in those regions^38–40^. Third-generation sequencing technologies (i.e., long-read) are excellent at tracking ARG hosts in environmental samples^28,41^ because long-read sequencing can directly reveal the genetic context of ARGs thanks to the extended read length. For example, two wastewater metagenomic studies reported greater numbers of long reads generated via Oxford Nanopore Technology (ONT) than numbers of short-read assembled contigs, and an average long read length of 2-10 kbp, which was significantly longer than the average length of short-read assembled contigs^28,42^. However, because both long- and short-read metagenomic sequencing are untargeted methods, their ability to detect low abundance and rare ARGs is limited^43,44^. On the other hand, emerging targeted methods such as single cell fusion PCR methods, called epicPCR (Emulsion, Paired Isolation, and Concatenation PCR), can overcome sensitivity limitations. In epicPCR single cells are isolated and encapsulated in a polyacrylamide bead within which PCR takes place to fuse a target ARG with the 16S rRNA gene^45^. As a result, PCR amplifies the signal of the ARG and its associated host 16S DNA from the background environmental metagenome, improving detection sensitivity. No studies to date have directly compared ARG hosts detected using untargeted sequencing and targeted fusion PCR methods.

In this study, we compared the sensitivity and consistency of ARG hosts detection via three different methods: short-read sequencing, long-read sequencing, and epicPCR. In addition, for the untargeted metagenomic sequencing methods, we also analyzed the genetic context of the detected ARGs, including their associations with MGEs. We then applied these methods to samples collected across a WWTP to characterize ARG hosts shifts across the wastewater treatment process, and to identify high-risk ARGs associated with putative pathogens and MGEs in the final effluent. The results of this work reveal breadth vs. sensitivity tradeoffs associated with method selection for identifying ARG hosts in wastewater monitoring programs.

## 2. Materials and Methods

### 2.1. Sample collection, DNA extraction, and pretreatment for epicPCR

Wastewater samples were collected from a conventional WWTP (City of West University Place WWTP, Houston, Texas, USA) that treats an average of 2 million gallons of municipal sewage per day. This WWTP employs a conventional aerobic activated sludge process as secondary treatment, followed by chlorination disinfection (gaseous Cl_2_, 2-4 mg/L effective chlorine concentration, 20 mins contact time). Nine grab samples were collected from three sampling locations, WWTP influent, secondary effluent, and final effluent on three consecutive dry days (n=3 for each sampling location). All samples were collected at the same time of the day to avoid diurnal variations. After collection, samples were kept on ice, immediately transported to the lab, and processed within 45 minutes of collection.

DNA was extracted from all samples prior to conducting long- and short-read metagenomic sequencing. A 50 mL influent sample, 250 mL secondary effluent sample, and 500 mL final effluent sample were filtered through a cellulose nitrate membrane filter (pore size 0.22 μm, diameter 47 mm; Millipore Sigma) to concentrate biomass. Next, filters were cut into small pieces using sterilized forceps and transferred to a 2 mL tube containing 0.1 mL glass beads for bead-beating, followed by DNA extraction. A Maxwell RSC Instrument (Cat. Num. AS4500, Promega) using Maxwell RSC PureFood GMO and Authentication kits (Cat. Num. AS1600, Promega) were used to extract DNA. For epicPCR, all influent (n=3) and final effluent samples (n=3) were centrifuged to concentrate cells for cell counting, polymerization, and cell lysis as previously described^85^. Only samples with good cell separation and partitioning in polyacrylamide beads (i.e., one single cell per 35-50 polyacrylamide beads) were used in the downstream experiments to avoid false positive detections. Details of DNA extraction and sample pretreatment for epicPCR are provided in Supplementary Information Section 1.2.

### 2.2. Sequencing epicPCR product using MinION (ONT)

We selected three ARG targets for epicPCR analysis: *sul1*, *ermB*, and *tetO*. They were chosen because of their wide host range as previously reported^24,86,87^. For example, *ermB*, the macrolide-lincosamide-streptogramin B (MLSB) resistance gene, is of clinical relevance because it is enriched in human-related environments, harbored by human pathogens, and often carried on MGEs^24^. Primer sequences for the three targets used in this study are listed in Supplementary Information Table 1. Details of the epicPCR experiments consisting of fusion PCR and nested PCR are provided in Supplementary Information 1.2. After attaining nested PCR products, library preparation and sequencing were performed following the protocol “Native barcoding amplicons (with EXP-NBD104, EXP-NBD114, and SQK-LSK 109)” (ONT). The pooled library was loaded on an R9.4 flow cell (MIN-FLO106, ONT) in a MinION device. The sequencing run was monitored via the software MinKNOW (v.20.10), targeting a >1000X depth per sample.

### 2.3. Metagenomics sequencing (long- and short-read)

DNA extracts of all samples were measured using a Qubit Broad Range dsDNA assay kit and Qubit 2.0 fluorometer. DNA quality was then evaluated using electrophoresis to ensure a DNA size greater than 3 kbps. For short-read sequencing, DNA extracts were shipped on dry ice to BGI Tech Solutions (Hong Kong) Co., Ltd for DNBseq general DNA library construction and DNBseq platform sequencing. For long-read sequencing, 500 ng of DNA from each of the three sample replicates were combined for library preparation following the protocol “Genomic DNA by ligation (SQK-LSK 109)” (ONT). Each of the three libraries (influent, secondary effluent, and final effluent) was loaded onto a Flow Cell R9.4 (MIN-FLO106, ONT) and sequenced with a MinION device. The sequencing run was controlled via MinKNOW (v.20.10). Long- and short-read sequencing statistics are provided in Supplementary Information Table 2.

### 2.4. Analysis of epicPCR reads for ARG host range profiling

Raw reads were basecalled via guppy_basecaller (Version 4.4.1+1c81d62). Basecalled reads were trimmed by Porechop (https://github.com/rrwick/Porechop) and filtered using Nanofilt with a minimum quality score of 7^88^. Next, all reads were searched against the corresponding linker primer sequence (RL-*sul1*-519F′, RL-*ermB*-519F′, and RL-*tetO*-519F′) using BLAST. The output reads were filtered using the perfect match criteria (100% identity and 100% length coverage) to exclude partially fused fragments, and only complete ARG-16S rRNA fusion structures were included for the downstream analysis. Then, we used a customized script to split the fusion structures into the ARG and the 16S rRNA gene portions based on the reverse linker position. We then conducted taxonomic classification on the 16S rRNA gene portion using Emu^89^ and the SILVA ribosomal RNA gene database (release 138, 2019). To avoid false positives, two actions were taken to further filter the reads: 1) The ARG portion of all split reads was aligned against the SARG database^90^ using BLAST, 2) for each sample, only hosts that were identified consistently from at least two sample replicates were counted. The results of epicPCR sequencing statistics can be found in Supplementary Information Section 2.1.

### 2.5. Analysis of metagenomic sequencing reads generated by long-read and short-read sequencing

We processed long- and short-read sequencing data in an integrated pipeline as shown in Supplementary Information Fig. 1. Our metagenomic analysis included: (1) identification of ARGs on long reads (via long-read sequencing) or short-read-assembled contigs (via short-read sequencing); (2) filter ARG-carrying long reads and contigs to include only those that were chromosome-associated for the host classification step; (3) identification of MGEs that were located on the same read or contig as the ARGs, and (4) identification of ARG host by taxonomic classification of the chromosomal reads or contigs that were associated with ARGs. Detailed methods describing the pipeline used to detect ARG-carrying long reads via BLAST, ARG-carrying contigs via CARD’s Resistance Gene Identifier (RGI)^91^, and ARG-MGE linkages are provided in Supplementary Information Table 7. ARG relative abundance was calculated by normalizing the copy number of ARGs detected on long reads or assembled contigs to the total giga base pairs (Gbp) of the sample^41^.

**Fig. 1.**
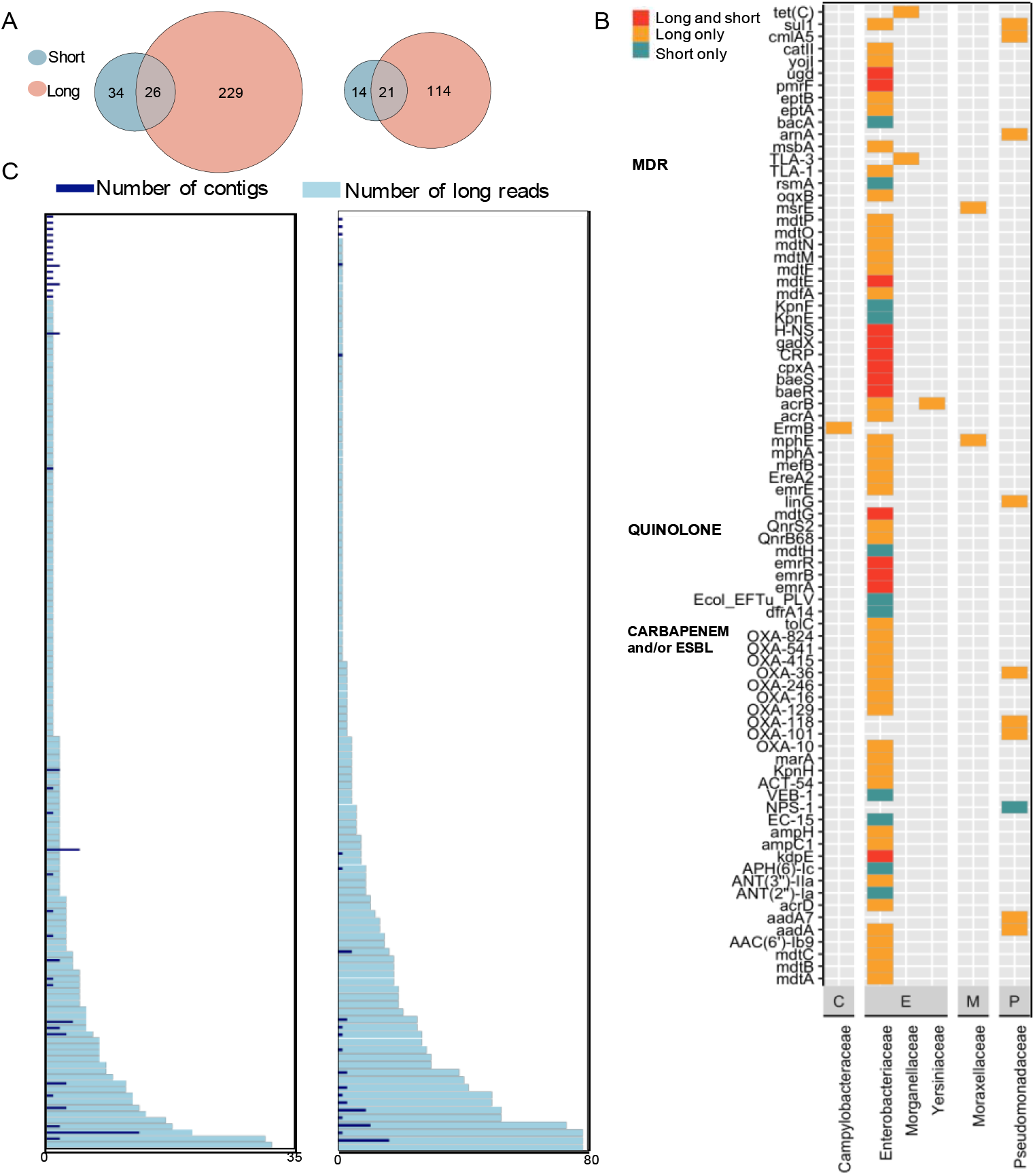
Comparison of long- and short-read sequencing for wastewater ARG host identification. **A.** Venn diagrams illustrating the detections of unique linkages of a specific ARG and its host family (left), and of unique linkages of a specific ARG drug class subtype and its host family (right). **B.** Profile of ARG bacterial hosts on the WHO priority list. Highlighted ARGs are those conferring multidrug resistance (MDR), fluoroquinolone resistance, and those encoding ESBL-production and/or carbapenemase-production. The colors denote a detection of an ARG-host (orange: detected only by long-read sequencing, turquoise: detected only by short-read sequencing, red: detected by both sequencing technologies). Family-level hosts are grouped by Order on x-axis (C: Campylobacterales, E: Enterobacterales, M: Moraxellales, P: Pseudomonadales). **C.** The number of reads (X-axis) via long-read sequencing (light blue bars) and the number of contigs via short-read sequencing (dark blue bars) supporting each unique linkage of ARG subtype and host family (Y-axis; specific linkages are not annotated on the graph). The left panel consists o data generated in this study, and the right panel is using publicly available data of a sample collected from the influent of a WWTP in Boston, MA (sample ID: B_ww_1, Supplementary Information Table 3).

ARG-carrying reads or contigs were categorized as “chromosome,” “plasmid,” or “unclassified” via PlasFlow (V1.1)^92^. The “unclassified” reads and contigs were re-classified via megaBLAST against the NCBI nt database with a minimum bit score of 50, an E value threshold of e^-10^, and a 70% sequence similarity cutoff, followed by keyword match (“chromosome”) to retrieve chromosome-associated long reads and contigs. All ARG-carrying reads and contigs classified as “chromosome” were subject to taxonomic classification using Centrifuge (V1.0.4)^93^. ARG-host linkages were identified by summarizing the associations between each ARG and the taxonomic classification result of the corresponding ARG-carrying read or contig. Putative pathogens were scanned according to the WHO resistant pathogen list^94^. In addition, three publicly available datasets from NCBI SRA were downloaded, each consisting of long-read (via Nanopore) and short-read (via Illumina) data based on sequencing the same wastewater sample^28,31^. These datasets were run through identical pipelines for analyzing long- and short-read sequencing data as used in this study to identify ARG-host linkages. Details regarding the three datasets are provided in Supplementary Information Table 3. Plasmid-associated ARG-carrying reads and contigs were subject to plasmid mobility prediction using MOB-suite (v3.0.3)^95^ and MOBscan (https://castillo.dicom.unican.es/mobscan/). Furthermore, to compare long-read sequencing with epicPCR for ARG host profiling, we processed Centrifuge specifically for those long reads that were found to carry *ermB*, *sul1*, and *tetO*.

## 3. Results and Discussion

### 3.1. Long-read sequencing demonstrated superior performance for ARG host identification as compared to short-read sequencing

Long-read sequencing identified a greater number of linkages of ARGs and their hosts than short-read sequencing (Fig. 1a). This result highlights that long-read sequencing produced a more diverse host profile than short-read sequencing, even though both methods produced similar total community bacterial composition profiles (Supplementary Information Table 4) and consistent ARG subtype profiles (Supplementary Information Fig. 2a). However, these two methods showed inconsistency in ARG host identifications. In total, 26 ARG-host family linkages, or 21 ARG subtype-host family linkages, were consistently detected by both methods, which accounts for only a small fraction of the corresponding total linkages detected by each method (Fig. 1a). Although several studies have focused on the consistency of long-read and short-read sequencing in resistome analysis^28,41,46^ and sample-wise taxonomic abundance estimation^47–50^, ours is the first to explicitly compare their ability to characterize ARG host range in wastewater and reveals inconsistencies across the methods (discussed below).

**Fig. 2.**
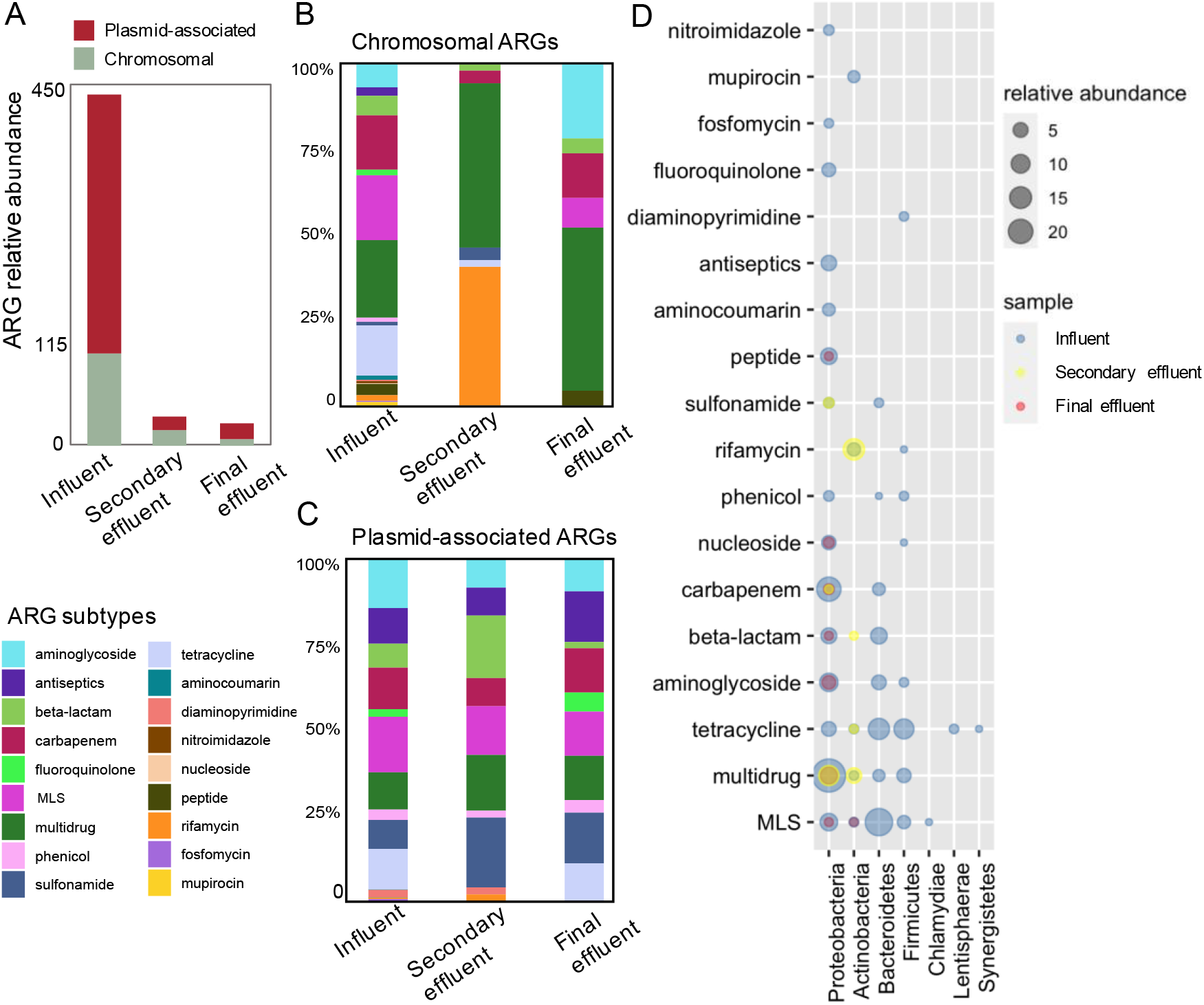
Dynamics of resistomes and ARG hosts across the WWTP revealed by long read sequencing. A. The relative abundance of ARGs across the WWTP (influent, secondary effluent and final effluent, n = 3 for each sampling location). ARGs were grouped by their location (red: plasmids, green: chromosome). B. The composition of chromosomal ARGs across samples. ARGs are colored by drug class subtype. C. The composition of plasmid-associated ARGs across samples. ARGs are colored by drug class subtype. D. The ARG host phyla across samples. ARGs are grouped by subtype (y-axis) and hosts are grouped by phyla (x-axis). The size of dots represents the relative abundance of ARGs corresponding to the specific subtype and host phylum. Dot colors indicate sampling location.

We further compared the ARG-host linkages identified by each method, focusing specifically on those putative hosts included on the WHO’s resistant pathogen list. Long- and short-read sequencing altogether recovered 117 ARG-host linkages, covering 80 ARGs (corresponding to 17 subtypes) and 31 putative pathogenic species (Fig. 1b). Both methods identified *Escherichia coli* as the putative pathogenic host that carried the highest abundance of ARGs. In fact, according to global surveillance of clinical cases, among all bacterial pathogens associated with or attributable to antibiotic resistance, *E. coli* ranks first as the cause of direct or indirect deaths^1^. Not surprisingly, most putative hosts identified were within the family *Enterobacteriaceae,* which includes the vast majority of commensal and enteric bacteria that live in the gastrointestinal tract of humans^51–53^. Consistent with previous studies, *Enterobacteriaceae* was found to harbor multiple classes of clinically relevant ARGs, especially those encoding ESBL (extended-spectrum beta-lactamase)-production and/or carbapenemase-production^51,54–56^. Long-read sequencing detected ARG-host connections across six host families and 68 ARGs, whereas short-read sequencing only detected *Enterobacteriaceae* and *Pseudomonadaceae* as the host families for 24 ARGs (Fig. 1b). Hence, despite having a significantly shallower sequencing depth (long-read method sequenced only 10.12% of the total bases sequenced by short-read method for the same wastewater sample; Supplementary Information Table 2), long-read sequencing detected a more comprehensive profile of putative pathogenic hosts of ARGs than short-read sequencing (Supplementary Information Table 5).

To investigate the inconsistency between the two sequencing methods, we compared the number of reads supporting the ARG-host linkages detected by long-read sequencing and the number of contigs supporting the ARG-host linkages detected by short-read sequencing (Fig. 1c). In addition, we also compared our results with an existing publicly available dataset containing short- and long-read metagenomic sequencing data of the same wastewater microbial community (ID: B_ww_1^31^; Fig. 1c, right; Supplementary Information Fig. 3). As expected, long-read sequencing demonstrated more ARG subtype-host family linkages as compared to short-read sequencing. Quantitatively, the numbers of long reads were generally greater than the numbers of contigs across the vast majority of the ARG subtype-host family linkages (Fig. 1c). For those reads and contigs that supported the same linkages, their numbers were moderately correlated (n=21, Spearman’s Rho=0.43, p<0.05 for this study; n=18, Spearman’s Rho=0.47, p<0.05 for B_ww_1), which indicates some degree of consistency between these two methods in quantifying the linkages of ARG subtypes and host families.

**Fig. 3.**
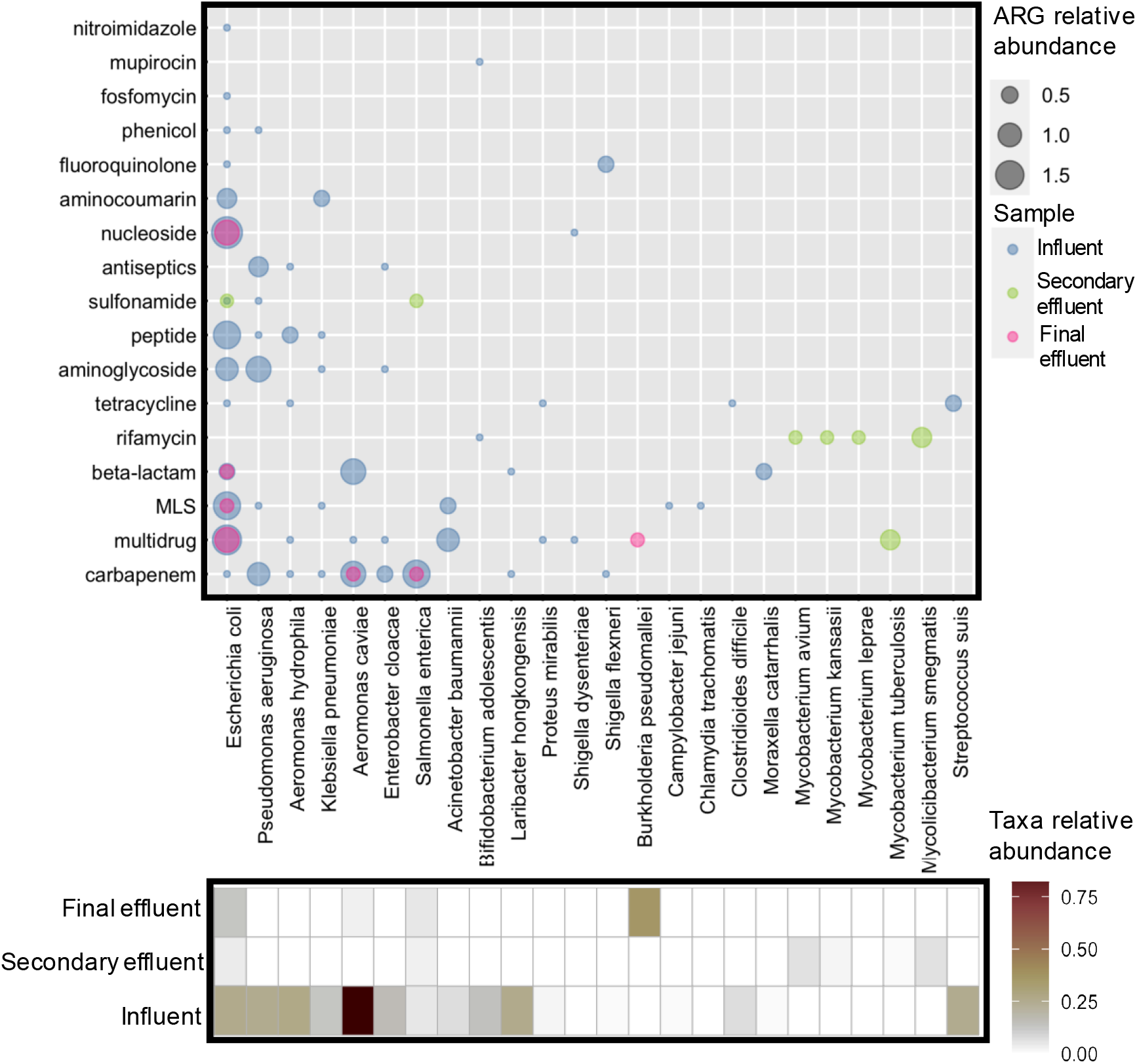
ARG-carrying putative pathogens detected in the influent, secondary effluent, and final effluent. ARGs are grouped by drug class subtype on y-axis; pathogenic species are shown on the x-axis. Dot size indicates the relative abundance of ARGs. Dot color indicates the sample location. The heatmap shows the taxa relative abundance of pathogenic species in each sample.

Thus, our results show that long-read sequencing demonstrated superior performance over short-read sequencing in detecting ARG hosts in two respects: 1) it captured a wider host range for different ARGs (Fig. 1b) and ARG subtypes (Fig. 1c, Supplementary Information Fig. 1a, 4); and 2) quantitatively, it detected ARG-host linkages by generating greater numbers of reads supporting the linkages as compared to short-read derived contigs (Fig. 1c, Supplementary Information Fig. 3). Of note, the evaluation was based on comparing the detection via long reads with the detection via contigs assembled from short reads, rather than directly comparing the raw reads generated by both sequencing methods. Several previous studies used raw short reads without assembly and identified putative ARG hosts through correlation analysis that compared the abundance of ARGs and taxonomical markers^57,58^. However, using raw reads to assign ARG hosts for wastewater surveillance has several challenges. First, this approach relies heavily on statistical correlation analysis which requires multiple sample replicates. Obtaining and processing multiple replicate samples significantly increases the amount of work required for sample collection, preparation, and sequencing, as well as the time and cost involved in routine surveillance. Most importantly, the raw read approach is prone to introduce false positives when detecting ARG-host linkages^59^. Thus, while using assembly is a relatively conservative means to identify ARG-host linkages as compared to using raw short-reads, it is less likely to generate false positives, which is crucial in wastewater surveillance and risk assessment. Furthermore, the substantial processing time and computational memory requirements of read assembly of short-read sequencing data can be massively reduced by using long-reads. Recent studies have shown that ONT, one of the leading long-read sequencing technologies, can rapidly and reliably detect resistomes and pathogens in one hour in wastewater^50^.

### 3.2. EpicPCR detected more ARG hosts as compared to long-read sequencing

Since long-read sequencing revealed more ARG hosts than short-read sequencing, we next compared the ARG hosts detected using long-read sequencing to the host range of three ARG targets (*sul1*, *ermB*, and *tetO*) detected by epicPCR. We found that epicPCR detected a greater number of host species for the three ARG targets than long-read sequencing (Table 1). As expected, epicPCR was more sensitive than long-read sequencing for ARG host detection as it is a targeted method that includes a PCR amplification step that enhances the signal of the target ARGs and bacterial host marker genes. An additional reason why epicPCR may be more sensitive than metagenomic sequencing is because it includes plasmid-associated linkages that may have been overlooked by metagenomics. With epicPCR, as long as the target ARG is present in the cell, it can be fused with the taxonomical marker (i.e., 16S rRNA gene) via PCR for host classification regardless of whether the ARG is located on a plasmid or chromosome. However, metagenomics analysis pipelines generally only classify hosts for ARGs that are located on the chromosome because the analysis requires the presence of taxonomic markers co-located on the ARG read, which are more frequently found in the chromosome than on plasmids. Thus, epicPCR should generate a more comprehensive host profile than metagenomics, because ARGs are widely distributed on plasmids^28,42,60^.

**Table 1.** The number of ARG-associated reads and host species detected by epicPCR and long-read sequencing in WWTP influent and effluent samples (n=3). The comprehensive list of hosts detected by epicPCR can be found in Supplementary Information Table 6.

| WWTP influent |  |  |  |  |  |  |
| --- | --- | --- | --- | --- | --- | --- |
| Method | EpicPCR | Long-read metagenomics |  |  |  | Both |
|  | Host species | Total reads | Host species via classifying total reads | Chromosomal reads | Host species via classifying chromosomal reads | Host species |
| *sulI* | 311 | 124 | 32 | 3 | 3 | 2 |
| *ermB* | 57 | 18 | 8 | 3 | 1 | 1 |
| *tetO* | 104 | 30 | 20 | 8 | 4 | 4 |
| WWTP effluent |  |  |  |  |  |  |
| Method | EpicPCR | Long-read metagenomics |  |  |  | Both |
|  | Host species | Total reads | Host species via classifying total reads | Chromosomal reads | Host species via classifying chromosomal reads | Host species |
| *sulI* | 323 | 8 | 7 | 0 | - | - |
| *ermB* | 14 | 0 | - | 0 | - | - |
| *tetO* | 35 | 0 | - | 0 | - | - |

The significantly lower number of hosts identified via long-read sequencing as compared to epicPCR was likely due to the low fraction of chromosomal reads among the total ARG-associated reads. To further investigate, we selected all long reads that were found to carry *sul1*, *ermB*, and *tetO* disregarding whether they were on chromosomes. As expected, the vast majority of ARG-carrying reads were not classified as chromosomal reads for the three ARG targets (Table 1). Unfortunately, it is not feasible to classify hosts using metagenomics for non-chromosomal ARGs, such as plasmid-associated ARGs. This is because phylogenetic analysis of plasmids is extremely challenging, due to the hardship to reconstruct the potentially shared “core” genes^61^ by plasmid subtypes^62,63^. In addition, plasmids can be horizontally transferred among different hosts and the transfer pattern is still not fully unveiled.

Despite epicPCR’s improved detection sensitivity over long-read sequencing, one undeniable value of long-read sequencing is its ability to capture the genetic context of an ARG. For instance, long-read sequencing identified the associations between *ermB* and the gene encoding conjugative transposon proteins in *Clostridioides difficile* (data not shown), highlighting *ermB*’s potential to be horizontally transferred. EpicPCR results in only a short, fused product of the target gene and taxonomic marker gene, retaining no additional contextual information in the sequenced amplicons. Thus, there is a tradeoff between sensitivity and contextual information that should be considered when deciding whether to use epicPCR versus long-read sequencing for ARG detection and risk assessment in environmental monitoring.

### 3.3. The ARG-host phylum linkages were relatively consistent across WWTP influent and effluent, whereas new ARG-host species linkages appeared in the WWTP effluent

ARGs were efficiently removed via conventional activated sludge treatment followed by chlorination disinfection, where we observed a 93.6% removal rate of all ARGs based on the relative abundance of total ARGs across the WWTP (Fig. 2a). The removal rate across the activated sludge treatment process (91.8%) was comparable to those reported in previous studies^14,30,64^. The chlorination process further removed chromosomal ARGs, but resulted in a slight increase in the relative abundance of plasmid-associated ARGs, leading to limited removal of total ARG relative abundance (21.0%) from the secondary effluent (Fig. 2a). Although the role of chlorination remains under debate with respect to its impact on antibiotic resistance^13^, several studies have shown that chlorination has a limited or even negative effect on the removal of ARGs from secondary effluent^19,65–67^.

To further understand the dynamics of resistomes across the treatment processes, the composition of ARGs on chromosomes and plasmids was assessed separately with respect to ARG subtypes (Fig. 2b). Chromosomal and plasmid-associated ARGs shared 16 ARG subtypes, whereas ARGs encoding resistance to fosfomycin and mupirocin were only detected on chromosomes, and ARGs conferring resistance to colistin and glycopeptide were only found on plasmids. The distribution of ARGs across chromosomes and plasmids is likely influenced by their resistance mechanism. We observed that ARGs causing antibiotic inactivation, replacement, or protection of the antibiotic’s target, were more frequently associated with plasmids than chromosomes, while ARGs associated with efflux pumps were more frequently located on chromosomes (Supplementary Information Fig. 4). This distribution pattern is consistent with a recent study investigating the distribution of ARGs across chromosomes and plasmids in major groups of *Enterobacteriaceae*^68^.

In general, the ARG subtypes present in secondary effluent and final effluent samples were a subset of those in influent samples (Fig. 2b,c). However, chromosomal ARGs (Fig. 2b) demonstrated a less consistent composition profile across the treatment processes as compared to plasmid-associated ARGs (Fig. 2c). Among the chromosomal ARGs, a spike of rifamycin resistance genes (i.e., *rpoB2*, *RbpA*, and *efpA*) in was observed in the secondary effluent and was likely attributed to the growth of their putative host Actinobacteria (Fig. 2d), whose relative abundance increased substantially in secondary effluent as compared to in the influent (data not shown). Similarly, the fraction of multidrug resistance genes (MDRs) increased in the secondary effluent (Fig. 2b), and these MDRs were also carried by Actinobacteria (Fig. 2d). The growth of Actinobacteria bacteria, which are common aerobes^69^, was likely the result of the presence of high dissolved oxygen concentrations in the activated sludge treatment process. In contrast, the relative abundance of the obligate anaerobic Bacteroidetes and the facultatively anaerobic Firmicutes decreased in the secondary effluent (Supplementary Information Table 5). Consequently, associations between Bacteroidetes or Firmicutes bacteria with ARGs were only observed in the influent samples (Fig. 2d). Proteobacteria was the dominant host phylum for ARGs across the entire wastewater treatment process (Fig. 2d), which is consistent with previous studies^70–72^. Our results indicate that the shift in the microbial community, and specifically the growth and decay of certain phyla drove changes in the resistome across the WWTP (Fig. 2b,c).

WWTP influent and effluent hosts were similar at the phylum level, as shown by both epicPCR and long-read sequencing (Supplementary Information Table 6 & Fig. 5), which is consistent with another study that used Nanopore sequencing for ARG host detection in WWTP influent and activated sludge^42^. However, at the species level, ARG hosts in the WWTP effluent were not entirely a subset of those in the WWTP influent due to the emergence of new hosts in the effluent (Supplementary Information Fig. 5). To gain a deeper understanding of the mechanisms responsible for the removal and selection of ARG hosts by different wastewater treatment unit processes, future research should focus on understanding the relative importance of horizontal gene transfer versus vertical propagation of ARGs via the growth and decay of ARG host, as well as the impact of environmental and operational variables on ARG propagation mechanisms^73^.

### 3.4. ARGs associated with pathogens and mobile genetic elements were present in the final effluent

We narrowed our focus to ARGs that were most likely to pose a public health risk using the following criteria: 1) presence in the final effluent, 2) association with an MGE, and 3) association with a pathogenic host species. Given the strong performance of long-read sequencing (i.e., high detection sensitivity on resistomes, hosts and MGEs), we performed this analysis using information obtained via long-read sequencing results. Notably, among all ARG-carrying reads in the effluent, 41.3% of them contained MDRs. ARGs associated with pathogens were abundant in the influent, which contained high abundances of enteric bacteria and as well as a diverse array of ARGs and ARG-carrying pathogens (Fig. 3).

We observed several ARG-carrying pathogens in the secondary effluent that were not detected in the influent, such as *Mycobacterium* species carrying rifamycin-resistance genes (Fig. 3).*Mycobacterium* is ubiquitous in wastewater and activated sludge and is considered a scavenger of insoluble compounds in wastewater^74,75^. One of the detected putative *Mycobacterium* pathogens, *Mycobacterium tuberculosis* (TB), was found to carry *efpA* which encodes an efflux pump system capable of extruding the isoniazid to the exterior of the cell^76^. This is particularly concerning because isoniazid is a drug commonly used in TB therapy. However, ARG-carrying *Mycobacterium* species were not found in the final effluent, indicating its effective removal in the disinfection process. *Burkholderia pseudomallei*, which can cause the disease melioidosis^77^, was an ARG host detected in the final effluent, but not in the influent or secondary effluent. It was associated with *MuxB*, a resistance-nodulation-cell division (RND) antibiotic efflux pump gene that significantly reduces susceptibility to macrolide, beta-lactams, and fluoroquinolones in bacteria.

Overall, six out of seven ARG-carrying putative pathogens present in the final effluent were also detected in the influent (Fig. 3). It is worth noting that the relative abundance of ARGs carried by *E. coli*, especially those encoding resistance against multidrug, beta-lactam, and nucleoside, were persistent across the entire treatment process (Fig. 3). In addition, the estimated relative abundance of total *E. coli* decreased significantly from influent to final effluent (Fig. 3). Together, these results suggest that the chlorination process may have selected for resistant *E. coli*. Previous studies have also observed that multidrug-resistant E. coli was persistent during wastewater treatment^78^ and was capable of escaping the oxidation by disinfectants^79^.

A variety of ARG-associated MGEs including *IntI1*s, recombinases, transposases, and integrases were frequently observed in the effluent samples (Supplementary Information Table 7), which suggests they may be involved in the HGT of ARGs among bacteria^80–82^. Recent studies revealed the striking prevalence of insertion sequences (IS) in resistant pathogens and the relatively consistent linkages between certain IS and specific ARGs across highly diverse bacterial genotypes, indicating the role of IS in mediating the HGT of these ARGs^83,84^. Similarly, we also found diverse IS families that were associated with ARGs across samples. Particularly, the IS6 family transposase was found to be frequently associated with two macrolide resistance genes across samples, namely, *msrE* and *mphE* (Supplementary Information Table 7). In addition, this specific *mphE*/*msrE*-the IS6 family transposase association was found on a conjugative plasmid equipped with the T4SS and MOBQ machinery, highlighting its potential for HGT via the interaction of IS and a conjugative plasmid.

## 4. Conclusions

The bacterial host and genetic context of an ARG present in our water and wastewater systems is critical to assessing its potential risk to human health. Specifically, ARGs of highest priority for further study are those hosted by pathogenic bacteria and/or with the potential to be horizontally transferred to pathogens (i.e., associated with an MGE). In this study, we evaluated and compared long and short-read metagenomics as well as epicPCR for identifying ARG hosts and associations with MGEs. We found that long-read sequencing outperformed short-read sequencing by generating a higher relative abundance of ARGs, especially of ARGs associated with MGEs, as well as a more diverse ARG host profile. Moreover, long-read sequencing generally yielded a greater number of reads supporting ARG-host linkages compared to the number of contigs assembled from short reads. EpicPCR outperformed long-read sequencing in terms of the breadth of hosts detected for three ARG targets (*ermB*, *sul1*, and *tetO*), however, it does not provide any additional contextual information (e.g., whether the ARG is associated with an MGE). When we applied these methods to understand ARG host dynamics across the WWTP, we observed consistent trends using long-read sequencing and epicPCR. Overall, the linkages of ARGs and host phyla in the WWTP effluent resembled those in the WWTP influent. However, at the species level, ARG hosts in the WWTP effluent were no longer a subset of those in the WWTP influent, which reinforces the need for more and longer-term surveillance of emerging effluent ARG hosts, and the importance of understanding the mechanisms of removal and selection of ARG hosts across treatment.

These results suggest that for environmental surveillance, long-read sequencing has many advantages as a tool for ARG detection and host tracking due to its high sequencing efficiency and because it does not require assembly. However, if any clinically-relevant ARG targets, such as MCRs (colistin resistance genes), are of particular concern to public health, epicPCR assays could be developed and applied to capture a more comprehensive host profile to complement routine metagenomic screening. Future studies should focus on evaluating standardized methods for wastewater-based surveillance of antibiotic resistance, developing guidelines for better reproducibility, and establishing a risk estimation framework for ARGs in the environment.

## Data Availability

All data produced in the present study are available upon reasonable request to the authors and sequencing data has been deposited to NCBI.

## Acknowledgments

This research was supported by funds from the National Institute of Food and Agriculture (grant no. 2016-68007-25044), the National Science Foundation (CBET 1805901 and 2029025, EF-2126387), a Johnson & Johnson WiSTEM2D 2D award, seed funds from Rice University, and a NIH grant from NIAID (P01-AI152999). We thank the operators at the West University City Place WWTP for helping with accessing wastewater samples.

## Data Availability

The metagenomic sequencing analysis pipeline and the epicPCR analysis pipeline are deposited at https://gitlab.com/treangenlab/wasterwater_arg_metagenomics. All sequencing data can be found at NCBI SRA (project accession number: PRJNA842493).

