## Supplementary materials for "Sensitivity and consistency of long- and short-read metagenomics and epicPCR for the detection of antibiotic resistance genes and their bacterial hosts in wastewater"

Supplementary Tables

Table 1. EpicPCR primers used for fusion PCR and nested PCR

Table 2. Long-read and short-read sequencing read statistics

Table 3. Accession numbers and brief descriptions of the three publicly available wastewater datasets used for comparing long- and short-read metagenomic sequencing

Table 4. Microbial community composition of the WWTP influent sample generated by long- and short-read sequencing

Table 5. ARG hosts detected by long-read sequencing, short-read sequencing, and epicPCR

Table 6. Hosts of *sul1*, *ermB* and *tetO* detected by long-read sequencing and epicPCR

Table 7. Associations between ARGs and MGEs detected by long-read sequencing

Supplementary Figure

Fig. 1. Overview of the computational pipeline for analyzing long- and short-read sequencing data

Fig. 2. Resistome profiles revealed by long- and short-read sequencing on paired wastewater samples

Fig. 3. Comparison of long- and short-read sequencing in identifying ARG subtypes-host family linkages for other publicly available datasets

Fig. 4. Composition of chromosomal ARGs and plasmid-associated ARGs in terms of resistance mechanisms across samples

Fig. 5. WWTP influent and effluent hosts revealed by epicPCR and long-read sequencing

### 1. Methods

#### 1.1 DNA extraction for long-read and short-read sequencing

After biomass concentration, filters were cut into small pieces using sterilized forceps and transferred to a 2 mL tube containing 0.1 mL glass beads for bead-beating. Immediately prior to bead-beating, 1 mL CTAB buffer was added to each sample tube. Sample tubes were vortexed for 30 seconds then incubated at 95°C for 5 minutes. Then, sample tubes were removed from heat and allowed to be cooled at room temperature for no more than 2 minutes. Next, sample tubes were bead beaten at max speed in a Mini-Beadbeater 24 (3,500 RPM; 112011, BioSpec) for 1 minute. After bead-beating, samples were briefly centrifuged and added with 40 μL proteinase K and 20 μL RNase A, then incubated at 70 °C for 10 minutes for lysis treatment. During the incubation, Maxwell RSC cartridges were setup following the manufacture's manual. A volume of 300 μL lysate from each sample tube was transferred to the cartridge. Each cartridge was also added with 300 μL lysis buffer. Finally, all cartridges with added sample lysate and reagents were loaded on to the instrument for automated extraction using the "Maxwell® RSC instrument with the PureFood GMO Protocol". After DNA extraction, DNA was eluted into 100 μL EB and stored in -80 °C before library preparations.

#### 1.2 EpicPCR

1.5 mL WWTP influent sample (n=3) was centrifuged at 10,000 g for 1 minute at 4 °C. 600 mL of final effluent sample (n=3) were centrifuged at 5,000 g for 10 minutes at 4 °C. Cell pellets were collected in a 2 mL microcentrifuge tube and washed in DNA grade ultrapure water for three times. Next, cells were agitated again using a vortex mixer at max speed (3000 RPM) for 45 seconds. Then, cells were diluted and stained with DAPI (4',6-diamidino-2-phenylindole) to perform cell count estimation on a Neubauer hemocytometer using a fluorescent microscope (IX 71, Olympus). A final cell count of 1 - 1.4 $\times 10^7$ in 30 µL of cell-water suspension was used for polyacrylamide bead formation and cell lysis treatment as previously described[1].

Next, fusion PCR and nested PCR were performed for each target per sample (n=3 each sample type). Fusion PCR was conducted within 24 hours after bead formation and cell lysis to prevent DNA degradation. For fusion PCR, PCR mastermix containing fusion templates (45 µL polyacrylamide bead solution), fusion PCR primers (the forward ARG primer F-*sul1*, F-*ermB* or F-*tetO*, the reverse 16S rRNA primer 1492R, and the linker primer RL-*sul1*-519F′, RL-*ermB*-519F′ or RL-*tetO*-519F′), Phusion HF DNA Polymerase (New England Biolabs), and emulsion stabilizers were homogenized in ABIL emulsion oil as previously described[2]. Fusion PCR conditions were optimized as follows: initial denaturation at 94 °C for 30 s; 35 cycles of denaturation at 94 °C for 5 s, primer annealing at 55 °C for 30 s, and extension at 72 °C for 30 s; a final extension step at 72 °C for 5 min. The primer sequences and amplicon sizes can be found in Table 1. Immediately after the fusion PCR reaction, 1 mM EDTA was added to the pooled sample, followed by diethyl ether/ethyl acetate wash[2]. Next, Monarch PCR & DNA Cleanup Kit (New England Biolabs) was used for DNA extraction from the washed beads. The purified DNA was eluted in 37 µL of EB and was subject to nested PCR. Next, nested PCR was performed using the purified fusion PCR products.

Nested PCR mastermix consisting of Phusion HF DNA polymerase, HF buffer, F’-*sul1*, F-*ermB* or F-*tetO,* and reverse 16S rRNA primer 1391R was divided into quadruplicate aliquots and combined with the purified fusion PCR products. The nested PCR program consisted of an initial denaturation for 30 s at 98 °C, followed by 38 cycles of denaturation at 98 °C for 5 s, primer annealing at 60 °C (for *sul1*) or 58 °C (for *ermB*) or 55 °C (for *tetO*) for 30 s, extension at

72 °C for 45 s, and a final extension step at 72 °C for 10 min. The final PCR products were loaded onto a 1.5% TAE agarose gel to confirm the expected product via electrophoresis (125 V, 50 minutes). DNA products of approximately 1 kbps in size were extracted using a Monarch gel extraction kit (New England Biolabs). The nested PCR products were purified again using AMPure XP beads and subject to library preparation.

**1.3 Integrated pipeline for analyzing long-read and short-read sequencing data**

We processed long-read and short-read sequencing reads in an integrated pipeline as shown in Fig. 1.

**1.3.1 Detections of ARG-carrying reads and ARG-carrying contigs, and the associations between ARGs and MGEs**

Long-read sequencing reads were screened for ARGs against the CARD database (V3.2.2) using BLAST (https://blast.ncbi.nlm.nih.gov) with a threshold of 70% identity and 70% length coverage. Short-read sequencing reads were assembled and processed using the resistance gene identifier (RGI, version 5.2.1[3]). Only contigs carrying the ARGs for which RGI produced "perfect" or "strict" match were selected for further analysis. To explicitly compare long-read and short-read sequencing on resistome characterization, ARG copy numbers generated by both methods were normalized against sequencing depth as previously described[4,5] to attain the relative ARG abundance in reads per billion bases sequenced (RPB). Classification of ARGs was conducted based on the oncology index file curated by CARD database. ARGs corresponding to at least two drug classes were classified as "multidrug" subtype. Beta-lactam resistant ARGs which confer resistance to carbapenem were selected and categorized as the "carbapenem" subtype.

ARG-carrying reads (long-read sequencing) and contigs (short-read sequencing) were subject to BLASTX under the minimum E-value $1\times10^{-5}$ with a length and identity threshold of 70% using a MGE database curated by NanoARG [4]. Long-read and short-read sequencing read statistics are provided in Table 2.

To evaluate the results of the comparison between long- and short-read sequencing in terms of ARG-host identification, we downloaded three publicly available datasets from two previous studies[6,7]. Both studies conducted long-read and short-read sequencing technologies to sequence the same wastewater samples. Details on the datasets are provided in Table 3.

##### 1.3.2 Sample-wise taxonomical abundance estimation for long-read and short-read sequencing data

The sample-wise taxonomical abundance estimation for long-read data was performed via Centrifuge v1.0.4. The program was run directly on ONT and Illumina reads and Centrifuge generated a report that contained the sample abundances.

#### 2. Results and discussion

##### 2.1 EpicPCR sequencing statistics

During read QC, we noticed that even though the size of PCR products was verified via electrophoresis, 50.94% of sequenced DNA still had a read length of shorter than 1 kbps, which may have been due to DNA fragmentation during gel purification and library preparation. We performed an alignment step during which reads were scrutinized for perfect match (i.e., 100% identity and 100% coverage) against the corresponding reverse linker primer sequence. This alignment step is the key to exclude false positives as it filtered out a substantial body of relatively short reads, which were likely partially fused PCR products. As a result, after this alignment step, the vast majority (71.8%) of remaining reads had a length falling within the range of 1007-1089 bps (i.e., the expected length range of nested PCR products given the primer design). Furthermore, the ARG portion of the remaining reads aligned to the corresponding ARG references in SARG database with relatively high sequence similarity and coverage. The average identity of alignment was 93.8 ± 3.1% for *ermB*, 93.9 ± 3.0 % for *sul1*, and 94.2 ± 3.0% for *tetO*. The average length coverage of alignment was 96.4 ± 9.8% for *ermB*, 99.4 ± 6.9 % for *sul1*, and 77.5 ± 5.3% for *tetO*. With respect to the 16S rRNA gene portion of reads, most remaining reads (91.8 ± 9.4%) passed the 16S rRNA gene alignment criteria of Emu (i.e., the 16S rRNA annotation tool used in this study)[8] and generated species-level classifications. These results underscore the successful acquisition of ARG-16S rRNA gene fusion structures.

#### 2.2 Direct comparison of long- and short-read sequencing for resistome analysis

##### 2.2.1 Long-read sequencing resulted in the detection of a more diverse and abundant resistome as compared to short-read sequencing

We first compared long- and short-read sequencing in their ability to characterize the diversity of ARGs (defined as the number of unique ARGs) and the relative abundance of ARGs (the copy number of ARGs normalized to sequencing depth) present in the samples. Overall, for raw wastewater (WWTP influent), long-read sequencing detected 347 ARGs with a total ARG relative abundance of 614 reads per billion bases (RPB), whereas short-read sequencing detected 191 ARGs with a total ARG relative abundance of 341 RPB. The total ARG relative abundance generated by both methods was comparable to previous metagenomic analyses that quantified ARGs of wastewater samples collected from western countries[9–11]. Therefore, in our study, long-read sequencing detected a significantly more diverse and abundant ARG profile as compared to short-read sequencing, which was surprising since the long-read sequencing depth was much shallower - only 10.12% of that of the short-read counterpart. One explanation for the better performance of long-read sequencing is its higher ARG detection sensitivity, underscored by the significantly higher proportion of ARG-associated reads among all long-read sequencing reads (0.0576%) as compared to the proportion of ARG-associated contigs among all short-read-assembled contigs (0.0119%).

The greater detection sensitivity of long-read sequencing is likely the result of a better preservation of the information of raw reads as compared to short-read sequencing. To elaborate, for short-read sequencing data, only 34.3% of raw reads mapped to the analyzed contigs, indicating a significant read loss during de novo assembly, which is a common issue for environmental metagenomes[12–14]. Of note, the analyzed contigs corresponded to those passed the length filter (1,500 bp), which accounted for approximately 23.1% of all assembled contigs, indicating the length of the assembled contigs was a limiting factor of ARG detection by short-read sequencing. In this study, short-read assembly was treated as a necessary step to avoid false positives caused by highly similar and relatively short ARG reference sequences. After de novo assembly, only 0.0125% of the assembled contigs passed through the filter of the ARG alignment step (based on RGI “perfect” and “strict” matches for ARG calling)[3]. For long-read processing, 100% of reads were directly subject to ARG alignment because no assembly was needed, and 14.8% of reads passed the filter of the ARG alignment step. Assembly of long reads was not performed due to the limited coverage (data not shown). Therefore, the number of ARG-carrying reads via long-read sequencing was greater than the number of ARG-carrying contigs via short-read sequencing (Table 2). Another reason long-read sequencing may have been more sensitive is because, on average, the size of the ARG-carrying reads (mean size=5,387 bp) was significantly larger than the ARG-carrying contigs (mean size=3,488 bp; $p=1.592e^{-05}$). Longer reads increased the likelihood of detecting multiple ARGs on the same reads. The number and fraction of long reads carrying more than one ARG (413; 23.7%) were greater than the number and fraction of contigs carrying more than one ARG (24; 10.7%). In addition, the contigs that carried more than one ARGs were carrying two or three ARGs, whereas 26.6% of the long reads that carried more than one ARGs were carrying at least three and up to six ARGs. Taken together, long-read sequencing resulted in higher ARG detection sensitivity than short-read sequencing by preventing read loss and through the generation of extended length of ARG-carrying reads.

However, both methods identified a comparable ARG composition with respect to ARG subtypes; each method detected the same suite of 20 ARG subtypes in wastewater (Fig. 2a). Approximately 90% of the total ARGs detected by each method belonged to subtypes of sulfonamide, macrolide-lincosamide-streptogramin (MLS), tetracycline, multidrug, carbapenem, aminoglycoside, antiseptics, and non-carbapenem-beta-lactams. The consistency between long-read and short-read sequencing in characterizing ARG subtypes has also been reported in other studies of wastewater samples and activated sludge samples[6], mock bacterial communities[15], and a plant population with a known bacteria spike[4].

##### 2.2.2 Long-read sequencing detected more ARGs located on chromosomes, plasmids, and on different types of mobile genetic elements (MGEs) as compared to short-read sequencing

Next, we compared the genetic location of ARGs assigned by each sequencing method. Both methods captured ARGs distributed across different genetic locations, namely, plasmid and chromosome. In addition, the associations between ARGs and MGEs (transposases, integrases, recombinases, and integrons) were also recovered. Long-read sequencing exhibited a greater abundance of ARGs that were associated with every single genetic location as compared to short-read sequencing (Fig. 2b). More specifically, long-read sequencing showed a significantly higher abundance of plasmid-associated ARGs, 6-fold higher than that of short-read sequencing, and a strikingly higher abundance of class 1 integron-integrase genes (*IntI1*)-associated ARGs, 16-fold higher than that of short-read sequencing (Fig. 2b). The less sensitive detection of MGE-associated ARGs by short-read sequencing was likely the result of the de novo assembly process. To elaborate, the variable copy number and the highly homologous and repetitive sequence compositions of MGEs make it problematic to assemble MGE-associated reads. As shown in a previous study, 82-94% of chromosomal sequences were correctly assembled and binned, but only 38-44% of genomic islands and 1-29% of plasmid sequences were identified in a simulated low-complexity short-read metagenome [16]. A similar degree of read loss during short-read assembly was also observed in several other studies of wastewater metagenomes[5,12,17,18]. Long-read sequencing, on the other hand, does not require assembly as it generates long reads that can be directly searched against MGE databases. Therefore, long-read sequencing can overcome the data loss issue associated with assembly, making it more feasible to detect ARG-MGE linkages.

So far, only a handful of studies have compared using long-read and short-read sequencing to determine the genetic locations of ARGs in wastewater samples. One study that obtained reads via Nanopore sequencing and contigs assembled from Illumina-sequencing reads found that both resulted plasmid-associated ARGs for all major subtypes of ARGs in wastewater and activated sludge samples[6]. Similarly, in this study, ARGs were found to be primarily located on plasmids rather than chromosomes (Fig. 2b). In addition, ARGs were mostly co-located with transposases and *IntI1* (Fig. 2b). We also investigated the distribution of ARGs across different genetic locations with respect to ARG subtypes (Fig. 2c). For ARGs conferring resistance to carbapenem, multidrug, MLS, diaminopyrimidine, aminoglycoside, tetracycline, nucleoside, and bacitracin, long-read sequencing demonstrated a consistent or slightly wider MGE distribution range compared to short-read sequencing (Fig. 2c). However, the distribution patterns of sulfonamide resistance genes, peptide resistance genes, rifamycin resistance genes, and antiseptics resistance genes were distinct for each method. Short-read sequencing assigned these ARGs only to plasmids whereas long-read sequencing assigned these ARGs not only to plasmids but also to other MGEs (Fig. 2c). This inconsistency was likely due to the significantly lower number of ARG-associated contigs detected by short-read sequencing for those specific ARGs (data not shown), which limited its ability to fully capture the potential of those ARGs being associated with MGEs. While this is not the first study to elucidate the genomic locations of ARGs by investigating the genetic context of ARGs, it is the first to explicitly compare long-read and short-read sequencing in profiling the distribution of ARGs across genomic locations (i.e., chromosomes, plasmids, and other MGEs) in wastewater.

**2.3 Host range detected by epicPCR**

For *sul1* hosts, epicPCR classified 61 Proteobacteria species and nine Bacteroidetes species (Table 5). NCBI Reference Sequence Database (RefSeq) reported consistent *sul1*-host phylum associations, more than 99% (25,195) of the reference sequences associated with *sul1* in bacteria were assigned to Proteobacteria. In previous studies characterizing hosts of *sul1* in wastewater, Bacteroidetes was identified as the dominant host phyla along with Proteobacteria[19,20]. To focus on the host range at the family level, the top three host families of *sul1* classified by epicPCR are Rhodocyclaceae, Aeromonadaceae, and Comamonadaceae, which was consistent with the *sul1* host range profiled by another targeted method using proximity ligation[21]. For *ermB* hosts, epicPCR identified 17 Proteobacteria species, 12 Bacteroidetes species, two Firmicute species, and one Fusobacteria species (Table 5). In RefSeq, *ermB* was predominantly associated with Proteobacteria (2,415 records), followed by Firmicutes (33 records). Recently, *ermB* has been found more frequently in Bacteroidetes species and was characterized as mobilizable based on its association with certain conjugative transposons[22,23]. Lastly, for *tetO* hosts, epicPCR detected 59 Firmicutes species, 6 Proteobacteria species, and 2 Bacteroidetes species. Consistently, according to NCBI RefSeq, *tetO* was found to be associated with Firmicutes (121 records), Proteobacteria (111 records), and Bacteroidetes (16 records). Almost all hosts detected by long-read sequencing were subsets of the host range detected by epicPCR as discussed in the manuscript. In addition, long-read sequencing and epicPCR demonstrated a consistent host range profile that *sul1* was mainly associated with Proteobacteria, *tetO* was mainly associated with Firmicutes, and *ermB* was mainly associated with Bacteroidetes and Firmicutes (Table 6).

**2.4 The profiles of ARG hosts across the WWTP influent and effluent revealed by long-read sequencing and epicPCR**

WWTP influent and effluent hosts were the most consistent at the phylum level as shown by epicPCR and long-read sequencing as discussed in the manuscript. In the WWTP effluent, as demonstrated by long-read sequencing, there were 12 ARG host species that were not detected in WWTP influent (hereafter referred to as "new hosts") as well as eight ARG host species which persisted across the whole treatment process (hereafter referred to as "persistent hosts"). In addition, none of the new hosts were found in the secondary effluent samples (data not shown). The persistent hosts included *E. coli* carrying *mdt* genes (i.e., *mdtE, mdtN,* and *mdtO*; subtype: efflux pumps) and *Aeromonas caviae* carrying *OXA-504* (subtype: multidrug/carbapenem). The new hosts included *Pseudomonas sp. BJP69* carrying *MexD* (subtype: efflux pump), Pseudomonas oleovorans carrying *mexF* (subtype: efflux pump), *Salmonella enterica* carrying *OXA-256* (subtype: multidrug/carbapenem), *Pandoraea thiooxydans* carrying *ceoB* (subtype: efflux pump), *Enterobacter kobei* carrying *ramA* (subtype: efflux pump), and *Burkholderia pseudomallei* carrying *MuxB* (subtype: efflux pump). Those ARG hosts (*E. coli, A. caviae, S. enterica, E. kobei, and B. pseudomallei*) are putative pathogenic species. This finding emphasizes a critical need to include them as the risk indicators, because they were harboring resistance genes of clinical relevance while at the same time poorly responsive to wastewater treatment.

Our results also showed that all ARG-carrying plasmids found in secondary effluent (containing 33 ARG-carrying plasmid reads) and final effluent (containing 56 ARG-carrying plasmid reads), as well as most (966 out of 970) ARG-carrying plasmids in influent, were classified as nonmobilizable plasmids due to the lack of a MOB. Only four out of 970 ARG-carrying plasmid reads in influent were found to carry MOB genes. However, although long-read sequencing generated greater length reads as compared to short-read assembled contigs, the average length of ARG-carrying, plasmid-associated reads was 4,885 bps. This suggests incomplete plasmids were assembled and thus may not have contained information needed to call mobility for a plasmid. For example, the length range of 14 representative ARG-bearing conjugative plasmids isolated from WWTPs was reported to be 35,925-290,014 kbps[24], much longer than the plasmid-associated read length. Therefore, we cannot draw a solid conclusion regarding the mobility of plasmids given that the relatively short plasmid-associated reads captured incomplete plasmid sequences.

Table 1. EpicPCR primers used for fusion PCR and nested PCR.

| Fusion PCR | | | |
|---|---|---|---|
| ARG | Primer | Sequence (5' - 3') | Reference |
| *sul1* | F-*sul1* | AAATGCTGCGAGTYGGMKCA | [25] |
| | RL-*sul1*-519F′ | GWATTACCGCGGCKGCTGAA CMACCAKCCTRCAGTCCG | [19] |
| *ermB* | F-*ermB* | GAACACTAGGGTTGTTCTTGC A | [26] |
| | RL-*ermB*-519F′ | GWATTACCGCGGCKGCTGCT GGAACATCTGTGGTATGGC | The reverse primer portion of *ermB*[26] |
| *tetO* | F-*tetO* | ACGGARAGTTTATTGTATACC | [27] |
| | RL-*tetO*-519F′ | GWATTACCGCGGCKGCTGTG GCGTATCTATAATGTTGAC | The reverse primer portion of *tetO*[27] |
| | 16S rRNA -1492R | GGTTACCTTGTTACGACTT | [1] |

| Nested PCR | | | | |
|---|---|---|---|---|
| ARG | Primer | Sequence (5' - 3') | Reference | Final product size (bp) |
| *sul1* | F’-*sul1* | GACGCCCTGTCCSRTCWGAT | [19] | 1037 |
| *ermB* | F-*ermB* | GAACACTAGGGTTGTTCTTGC A | [26] | 1007 |
| *tetO* | F-*tetO* | ACGGARAGTTTATTGTATACC | [27] | 1043 |
| | U519F-block10 | TTTTTTTTTTCAGCMGCCGCG GT AATWC/3SpC3/ | [1] | |
| | U519R-block10 | TTTTTTTTTTGWATTACCGCG GC KGCTG/3SpC3/ | | |
| | 16S rRNA -1391R | GACGGGCGGTGTGTRCA | [28] | |

Table 2. Long- and short-read sequencing read statistics.

| Method | Sample | Total bases | Number of reads/contigs | Length N50 (bp) |
|---|---|---|---|---|
| Long-read sequencing | Influent (n=3) | 4,318,719,998 | 1,178,106 | 4,748 |
| Short-read sequencing | | 42,663,854,700 | 1,796,758 | 1,210 |
| Long-read sequencing | Secondary effluent (n=3) | 2,885,452,624 | 464,436 | 7,159 |
| short-read sequencing | Final effluent (n=3) | 1,944,820,196 | 1,513,866 | 1,493 |

Table 3. Accession numbers and brief descriptions of the three publicly available wastewater datasets used for comparing long- and short-read metagenomic sequencing

| ID | Sequencing platform | Instrument | Total bases | Sample | Sampling region | SRR Accession | Reference |
|---|---|---|---|---|---|---|---|
| ST _IN | Illumina | Illumina HiSeq 4000 (PE 150) | 18G | Municipal wastewater | Hong Kong | SRR8208343 | 6 |
| | ONT | ONT MinION | 2.4G | | | SRR7497167 | |
| B_WW_1 | Illumina | Illumina HiSeq 2500 (PE150) | 5.9G | | The greater Boston area, in Massachusetts, USA | SRR12917052 | 7 |
| | ONT | ONT MinION | 1.3G | | | SRR12917048 | |
| B_WW_2 | Illumina | Illumina HiSeq 2500 (PE150) | 5.6G | | | SRR12917051 | |
| | ONT | ONT MinION | 0.82G | | | SRR12917047 | |

Table 4. Microbial community composition of the WWTP influent sample generated by long- and short-read sequencing

| Family | Relative abundance via long-read sequencing | Relative abundance via short-read sequencing |
|---|---|---|
| Enterobacteriaceae | 4.66 | 6.24 |
| Bacillaceae | 0.58 | 3.1 |
| Pseudomonadaceae | 3.22 | 3.08 |
| Streptomycetaceae | 1.03 | 2.22 |
| Flavobacteriaceae | 0.67 | 2.16 |
| Burkholderiaceae | 1.67 | 1.9 |
| Lactobacillaceae | 0.26 | 1.88 |
| Streptococcaceae | 1.22 | 1.73 |
| Mycobacteriaceae | 0.73 | 1.56 |
| Microbacteriaceae | 0.5 | 1.45 |
| Xanthomonadaceae | 1.05 | 1.35 |
| Rhizobiaceae | 0.63 | 1.23 |
| Corynebacteriaceae | 0.16 | 1.23 |
| Mycoplasmataceae | 0.11 | 1.22 |
| Sphingomonadaceae | 0.49 | 1.07 |
| Campylobacteraceae | 0.49 | 1.04 |
| Vibrionaceae | 0.39 | 1.03 |
| Paenibacillaceae | 0.35 | 0.98 |

| | | |
|---|---|---|
| Comamonadaceae | 5.06 | 0.97 |
| Pasteurellaceae | 0.45 | 0.97 |
| Staphylococcaceae | 0.16 | 0.95 |
| Moraxellaceae | 1.82 | 0.89 |
| Yersiniaceae | 0.32 | 0.89 |
| Clostridiaceae | 0.48 | 0.87 |
| Roseobacteraceae | 0.24 | 0.83 |
| Species | Relative abundance via long-read sequencing | Relative abundance via short-read sequencing |
| Salmonella enterica | 0.05 | 2.09 |
| Escherichia coli | 0.26 | 1.16 |
| Bacillus cereus group | 0.1 | 0.57 |
| Helicobacter pylori | 0.02 | 0.47 |
| Bacillus subtilis group | 0.04 | 0.44 |
| pseudomallei group | 0.11 | 0.42 |
| Listeria monocytogenes | 0.01 | 0.38 |
| Pseudomonas syringae group | 0.08 | 0.35 |
| spotted fever group | 0.01 | 0.29 |
| Enterobacter cloacae complex | 0.42 | 0.28 |
| Pseudomonas aeruginosa group | 1.08 | 0.27 |
| Burkholderia cepacia complex | 0.26 | 0.26 |
| Buchnera aphidicola | 0.03 | 0.26 |
| Staphylococcus aureus | 0.01 | 0.26 |
| Burkholderia pseudomallei | 0.04 | 0.25 |
| Yersinia pseudotuberculosis complex | 0.01 | 0.22 |
| Pseudomonas fluorescens group | 0.14 | 0.2 |
| Acinetobacter calcoaceticus/baumannii complex | 0.12 | 0.2 |
| Pseudomonas putida group | 0.17 | 0.19 |
| Klebsiella pneumoniae | 0.13 | 0.19 |
| Pseudomonas aeruginosa | 0.25 | 0.18 |
| Bacillus amyloliquefaciens group | 0.01 | 0.18 |
| Mycobacterium avium complex (MAC) | 0.03 | 0.17 |
| Bacillus thuringiensis | 0.01 | 0.17 |
| Vibrio harveyi group | 0.06 | 0.16 |

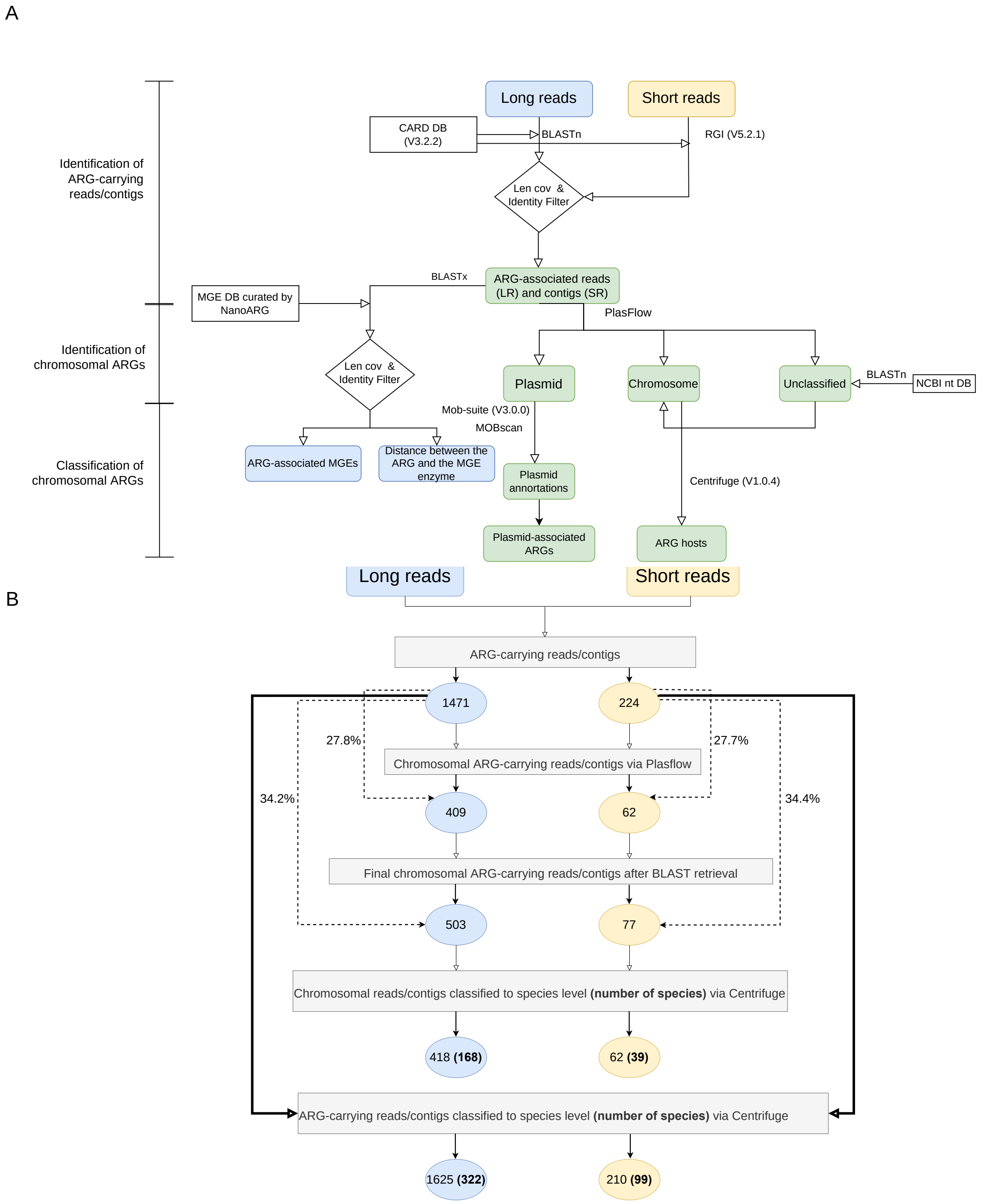


**Fig. 1.** Overview of the computational pipeline for analyzing long- and short-read sequencing data. A. The detailed pipeline. B. The output of each step, in terms of the number of reads (long-read sequencing) and contigs (short-read sequencing), and the number of host species classified based on chromosomal ARGs and all ARGs.

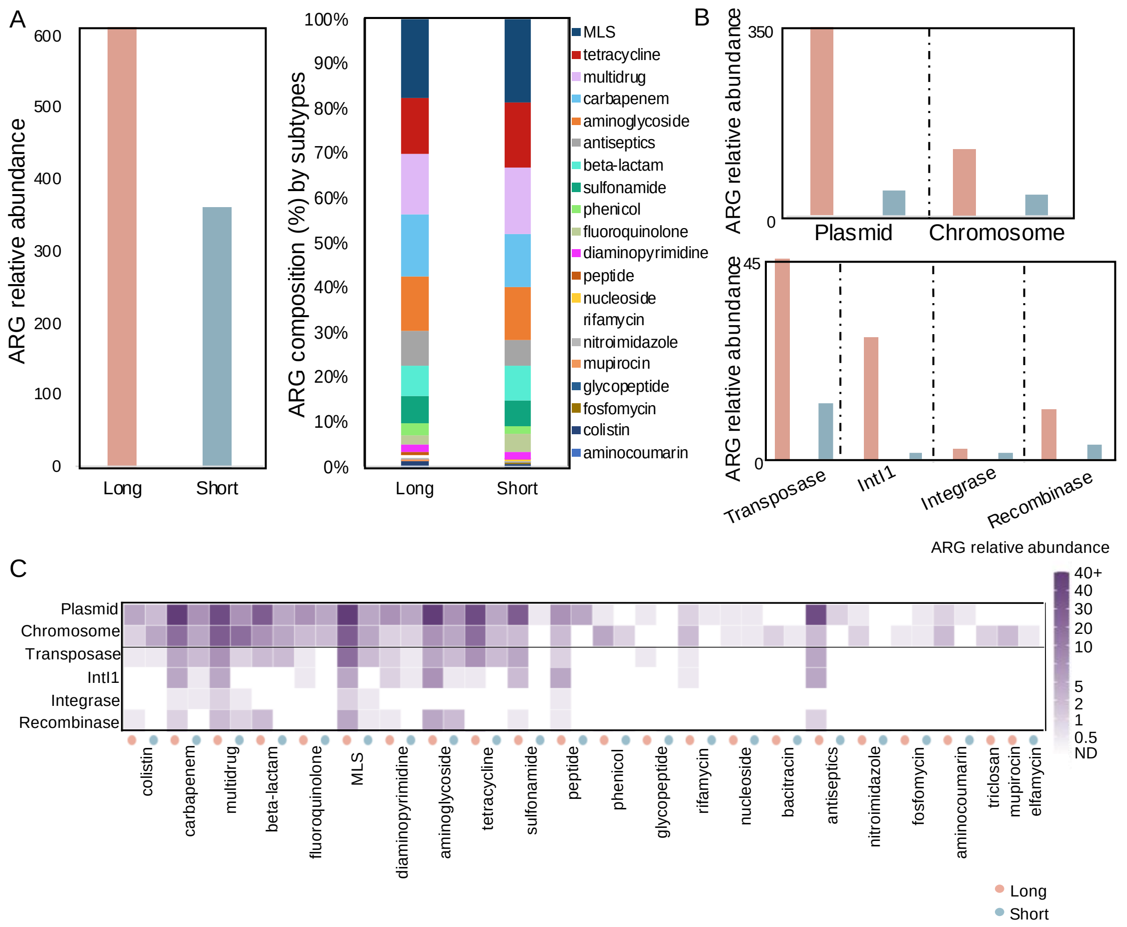


**Fig. 2**. Resistome profiles revealed by long- and short-read sequencing on paired wastewater samples (n = 3). A. Total ARG relative abundance revealed by long- and short-read sequencing (left) and ARG composition broken down by drug class subtype (right) according to the relative abundance of ARGs of each subtype. B. Distribution of total ARGs across genetic locations (plasmid or chromosome) and the associations between ARGs and MGEs as determined by long- and short-read sequencing. ARGs associated with more than one MGE were counted separately for each MGE involved. C. Distribution of ARGs (grouped by drug class subtype on the x-axis) across genetic locations and ARG-MGE associations revealed by long- and short-read sequencing.

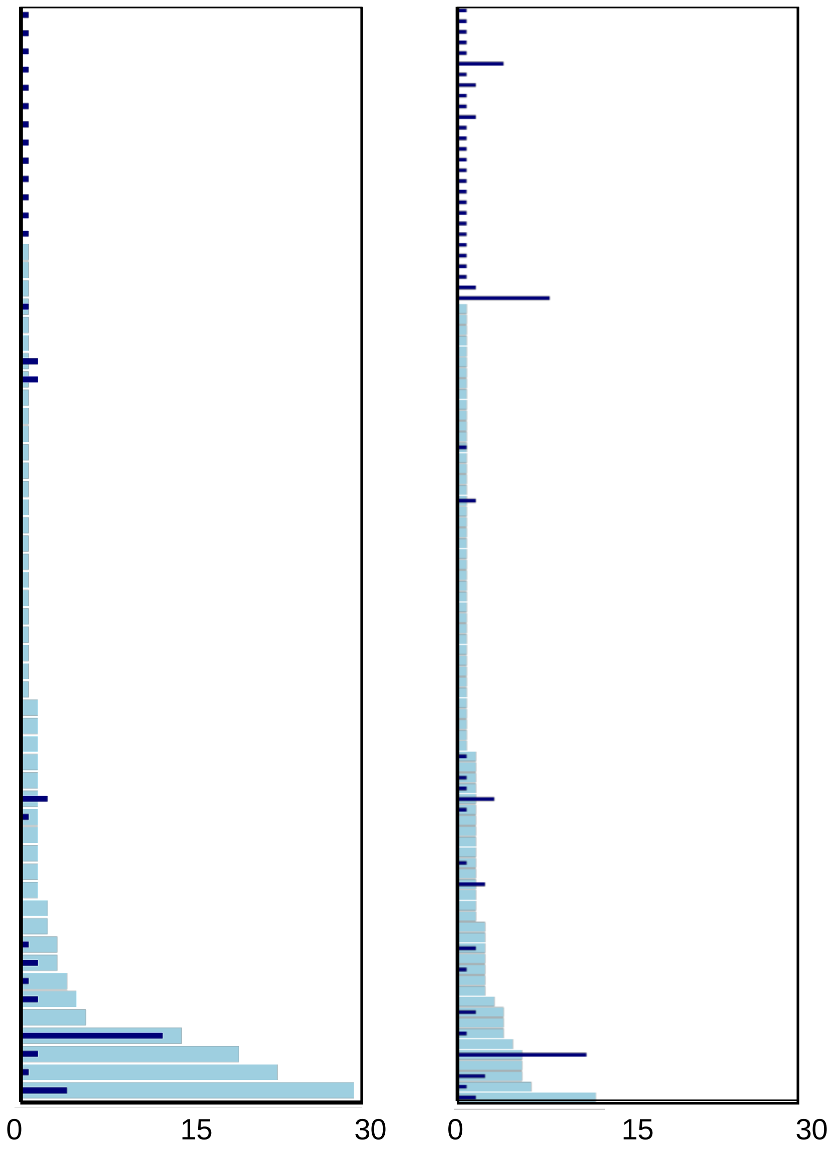


**Fig. 3.** Comparison of long- and short-read sequencing in identifying ARG subtypes-host family linkages for other publicly available datasets. Left: sample ID: B_WW_2[7], right: ST_IN[6].

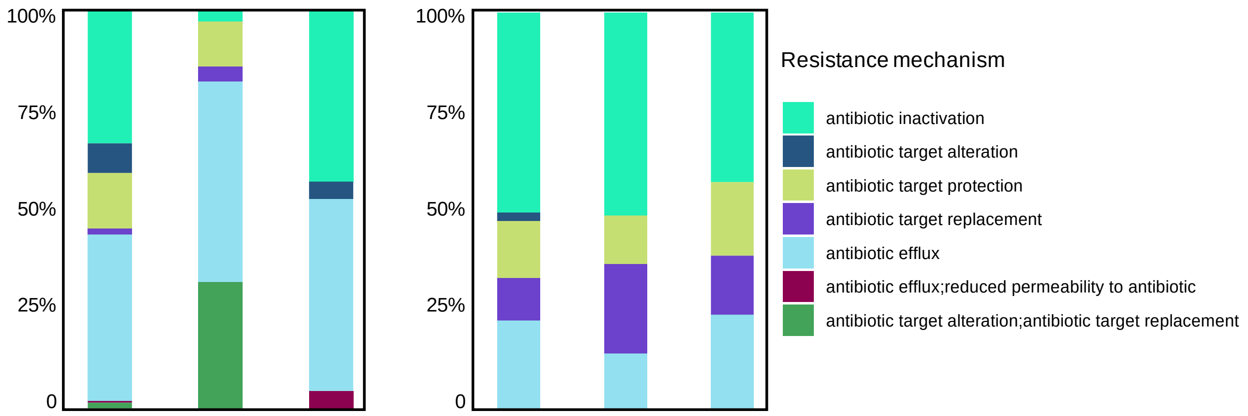


**Fig. 4.** Composition of chromosomal ARGs and plasmid-associated ARGs in terms of resistance mechanisms across samples.

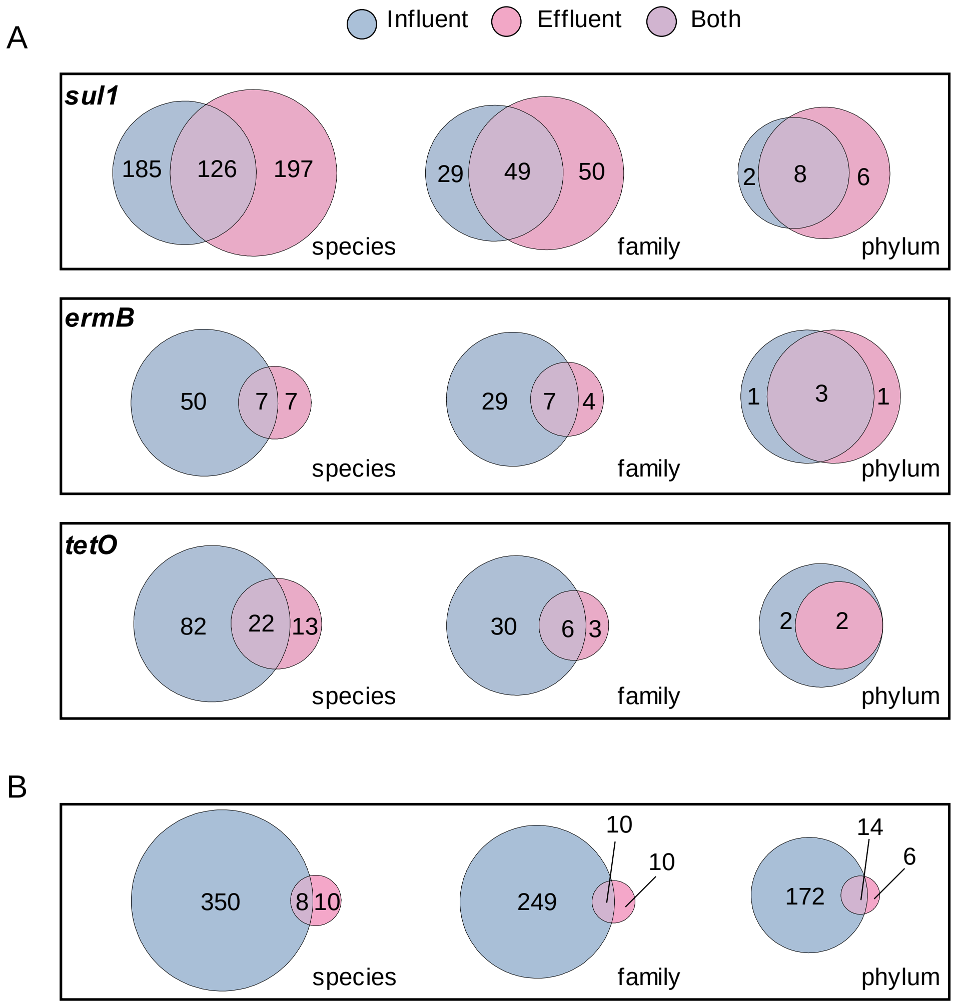


**Fig. 5.** WWTP influent and effluent hosts revealed by epicPCR and long read sequencing. a. The count of WWTP influent (blue) and effluent (violet) hosts revealed by epicPCR. The numbers of hosts for the three ARGs (*sul1*, *ermB* and *tetO*) are shown at species, family, and phylum level, respectively. b. The count of influent (blue) and effluent (violet) hosts revealed by long-read sequencing. Unique combinations of each ARG and its host counted at the species, family, and phylum level, respectively.
